# Evaluating Oxidative stress marker F_2α_-Isoprostanes relations to early life adversity and as a predictor of neurodevelopment in infants and toddlers

**DOI:** 10.1101/2025.10.20.25338379

**Authors:** Kameelah Gateau, Ramon Durazo-Arvizu, Pat Levitt

**Affiliations:** Division of Neonatology; Neurology; Department of Pediatrics, Keck School of Medicine of University of Southern California, Los Angeles, CA, USA; The Saban Research Institute, Children’s Hospital Los Angeles, Los Angeles, CA, USA

**Author notes:** Corresponding Author: Kameelah Gateau MD,MS.

**Keywords:** Early life adversity, biomarkers, isoprostanes, early childhood development

## Abstract

**Background:** Experiencing early life adversity (ELA) and chronic stress activation early in childhood increases the risk for altered developmental trajectories that lead to lifelong impacts on physical and mental health. This results in a toxic stress response – the dysregulation of neuroendocrine, immune and metabolic functions that causes allostatic load and a higher risk for poor health. A major challenge in current pediatric screening practices for identifying toxic stress is the typically use of parent retrospective or child prospective Adverse Childhood Experience (ACEs) questionnaires, which are no better than chance in predicting poor health outcomes. We have proposed that the adaptative processes to ELA converge on mitochondrial function and is measurable in several ways, including the gold standard marker of oxidative stress, F_2α_-Isoprostanes (F2_α_ - IsoPs). The aim of this study was to evaluate the relation between F_2α_-IsoPs measures and child neurodevelopment and how those relations change over critical developmental periods.

**Methods:** In our ongoing “Family First” longitudinal study, F_2α_-IsoPs were measured from child urine samples collected at 6-,12-, and 24 months. Maternal adversity was assessed using the ACES questionnaire at 6- and 12-months. The Bayley 4 was administered to assess neurodevelopment at all study time points.

**Results:** Maternal ACEs scores were significantly correlated to higher F_2α_-IsoPs levels at 12 months. Elevated infant F_2α_-IsoPs at 6 months were correlated significantly with lower language and motor scores Bayley scores. Assessment of the relation between maternal ACEs on changes in F_2α_-IsoPs levels over the first year of life demonstrated there were significantly different F_2α_-IsoPs trajectories for infants whose mothers endorsed 0, 1-2, or 3+ ACEs.

**Conclusions:** The data indicate that there are specific physiologic contributions of mitochondrial generated oxidative stress to a toxic stress response in young children. A broader approach to pediatric screening for toxic stress predictability may be the incorporation of F2_α_ -IsoPs measures in the first two years postnatal.

## Introduction

It has been over two and a half decades since the study by Fellitti et. al established the dose dependent relationship between experiencing early life adversity (ELA) (including but not limited to physical or psychological abuse by a caregiver and household dysfunction) and significant morbidity and mortality in adulthood(1). In the time since the original report, many population-based studies have shown that experiencing ELA is relatively common, with 63% of the US population having experienced at least 1 adverse childhood experience (ACE) and 22% with 4 or more ACEs. The latter category predicts, at a population level, a significantly increased risk for poor adult health, including cardiovascular disease, immune dysfunction, obesity and diabetes, pulmonary and mental illnesses(1-3). Studies have subsequently found that ACES may actually be more pervasive than previously identified when expanded domains of adversity are assessed, including household unpredictability, peer and community level violence, discrimination, and racism(4-7). When looking at the most critical stage of development in utero, studies also have found that perinatal and early postnatal adversity experienced by the mother can alter the developing fetal brain and postnatal child health, behavior, and neurodevelopment outcomes. These experiences forecast poorer health not only for the mother, but also intergenerationally for their offspring (8-14) . Current screening practices for adversity utilize retrospective self-report questionnaires that alone have been shown to have no better than chance at predicting individual health outcomes(15-17). Additionally, such screening does not provide physiologic measures that could assist in developing best practices intervention strategies. A 2024 paper written by 10 experts in the ELA field reported that despite there being substantial literature linking early life adversity to a toxic stress response, allostatic load, and poor health outcomes, there is no consensus guideline on how to screen or diagnose toxic stress response or allostatic load(18).

There has been emphasis on combining questionnaire-based screening with identifying and measuring the physiologic changes that result from experiences of ELA. McEwen and colleagues proposed a physiologic pathway of maladaptation that leads to allostasis and if chronic, a shift to allostatic load. The 2005 report from the National Scientific Council on the Developing Child introduced the term toxic stress, defined as the neuro-endocrine-immune systems dysfunction that results from prolonged and severe experiences of adversity, as the pathophysiologic changes that tip allostasis to allostatic load(19-24). Over time with continued exposure to adversity the allostatic load maladaptive state persists, even when the stressor is no longer present(10, 19, 23, 25-27). McEwen, Picard and colleagues proposed that the neuro-endocrine-immune systems dysfunctions are in part mediated through disrupted mitochondrial function. Mitochondria play a key role in adapting to changing energy demands during development along with adaptation and maladaptation to chronic stressors(28-35).

Reactive oxygen species (ROS) are normally generated as a part of cellular respiration and accumulate in excess when mitochondria malfunction. This leads to oxidative stress and eventually, cellular damage(36). Oxidative stress can been measured in children using F_2α_−Isoprostanes (F_2α_-IsoPs)(37-44). The Biomarkers of Oxidative Stress Study established F_2α_-IsoPs as the gold standard oxidative stress measure, generated via non-enzymatic, free-radical catalyzed peroxidation of arachidonic acid, serving as dose- and time-dependent markers of endogenous oxidative stress(39, 45). Oxidative stress and mitochondrial dysfunction have been studied extensively in adult disease and aging but lacking in developmental contexts(46-56).

The causal pathway linking chronic stress and adversity to poorer health as mediated by oxidative stress is supported by basic science and several human exploratory studies, reporting associations between ELA and higher oxidative stress in animal models, infants and adolescents (57-59). Studies have also found that prenatal adversity in mothers is correlated with elevated oxidative stress in their infant, and elevated oxidative stress in mothers is associated with social impairments in school age children(58, 60). Pre-clinical models from our group have also found that reducing oxidative stress in an ELA model mice normalized disrupted mitochondrial function and reward behavior(61).To our knowledge there have been no prospective longitudinal studies evaluating the relationship between maternal adversity, child oxidative stress and developmental outcomes. Here, the aim of this study is to identify the relation between maternal ELA, its potential impact on oxidative stress measures of their child. We also aim to identify how oxidative stress in infants in toddlers impacts neurodevelopment over their first 2 years of life.

## Methods

### Study participants

For this longitudinal observational study, maternal-infant dyads (340) were largely recruited (approximately 70%) from Kaiser-Permanente Medical Centers in Los Angeles County, with the other 30% of participants from local Los Angeles community clinics and hospitals and through social media. Inclusion criteria were maternal age 18 or older, gestational age between 37-42 weeks (full term) or 32-36 weeks (prematurity), and recruitment at 150 days postnatal or younger. Exclusion criteria were gestational age < 32 weeks, NICU stays > 7 days, severe sensory or motor impairments (e.g. blindness, deafness, cerebral palsy), identified metabolic, syndromic or progressive neurological disorders (e.g. Down Syndrome, epilepsy, Fragile X), perinatal complications requiring intubation, grade 1 or greater intraventricular hemorrhage, periventricular leukomalacia, congenital malformations, sibling with an autism diagnosis, multiples (twins or greater). Mothers were consented for all study timepoints and provided consent on behalf of their child. All study protocols were reviewed and approved by the Children’s Hospital Institutional Review Board.

#### Caregiver Questionnaires

##### Contact Information & Demographics Forms

This form asked participants to provide their home address, phone number, email, partner information and best times to contact, occupation, income, marital status, and living conditions.

##### Medical History Form

Information was collected from before and during pregnancy and current physical and mental health diagnoses of the mother, birth history of the infant, and infant measures (length, weight, head circumference) at birth. Immediate family members and sibling health information also was collected.

##### Perceived Stress Scale (PSS)

This is a 10-item questionnaire that queries mothers about feelings and stress during the past month(62). Scores range from 0-30 with scores less than 13 are categorized as low stress, scores 14-26 as moderate stress and scores 27 and above considered high stress.

##### Patient Health Questionnaire-9 (PHQ-9)

This questionnaire assesses depressive symptoms over the prior 7 days. Scores range from 0-30 with a score of 13 or greater indicating clinical depression(63).

##### Adverse Childhood Experiences Questionnaire for Adults (ACEs)

This is a standardized, retrospective questionnaire for adults, gathering information on childhood experiences that may have an impact on their health as an adult(1). Mothers are asked to report on past events happening in their childhood through age 18. The 10 specific questions are deidentified, yielding an aggregated score.

##### Questionnaire of Unpredictability in Childhood (QUIC-5)

This validated questionnaire consists of 5 questions and assesses unpredictability in the child and mother’s childhood environment based on a report of typical home experiences(64). It has validity for identifying risk of internalizing symptoms. In infants and children, unpredictable caregiver signals are associated with poorer memory, disrupted self-regulation and altered brain development(65, 66).

##### Pediatric ACEs and Related Life Events Screener (PEARLS)

Is a two-part questionnaire that collects information from the mother on the child’s exposure to the validated, standard adverse childhood experiences (part 1) (1)and other adversities and stressful life events(67). The screener is deidentified for individual items, yielding an aggregated score.

#### Direct Child Development Measures

##### The Bayley-4 Scales of Infant and Toddler Development (Bayley-4)

This direct measure of cognition, language and motor development is administered by trained and validated research staff. Cut scores are generated by age for each domain(68). Instructions are provided in English or Spanish at the request of the mother.

#### Oxidative stress

##### F_2α_-Isoprostanes

Infant urine collection is done using a diaper insert bag or a urine cup for older children. Samples (1-3ml aliquots) are frozen at -80°C within an hour of collection and shipped in bulk to the Eiconasoid Core Laboratory at Vanderbilt University, with duplicates kept at CHLA. F_2α_-IsoPs content is measured by gas chromatography/negative ion chemical ionization mass spectrometry using established methods(41, 44). The two most abundant regiosomers 15-F_2t_-IsoP and 5-F_2t_-IsoP are quantified and reported here. Results are standardized to creatinine to account for variations in renal function that may affect F_2α_-IsoPs concentrations in urine.

#### Statistical Analysis

Demographics were calculated as a percentage of all participants who responded. Adversity questionnaire percentages were calculated based on ACEs (0, 1-3, 4+) and PEARLS (0 or >0) categories. Mean and standard deviations were calculated for all oxidative stress measures and Bayley development scores. A mixed effects model was used to evaluate the change in F_2t_-IsoP between 6-24 months. Linear splines with a single knot at 6 months were used to estimate the non-linear time trend(69). Pearson correlation coefficients were used to calculate relations between maternal ACEs, QUIC5, Bayley 4 scores and F_2t_-IsoP levels. IBM SPSS Statistical Analysis Package version 29.0.0.0(241) and R version 4.4.1 were used for data analyses.

### Results

### Participant Demographics

In this ongoing longitudinal study, 340 maternal-infant dyads were enrolled. The study visits for the dyads occur when the child is in the time window for 6-,12-,24-,36-,48 months. The data presented here is inclusive up to the 24-month visit, as the later time point visits are at the early stages of occurring. Demographic information of the study participants is presented in Table 1. Notable features include an even distribution by sex (50.6% female, 49.4% male), racial classification that includes 50% of participants self-identifying as White and 64% of participants self-identifying as Hispanic/Latine (Table 1). Maternal educational attainment included 23% completing high school or GED, 28% a bachelor’s degree, and 22.6% a master’s degree (Table 1). Annual combined family income included 27.8% study participants reporting <$50,000 with the median income in Los Angeles being approximately $80,000(70).

**Table 1.**
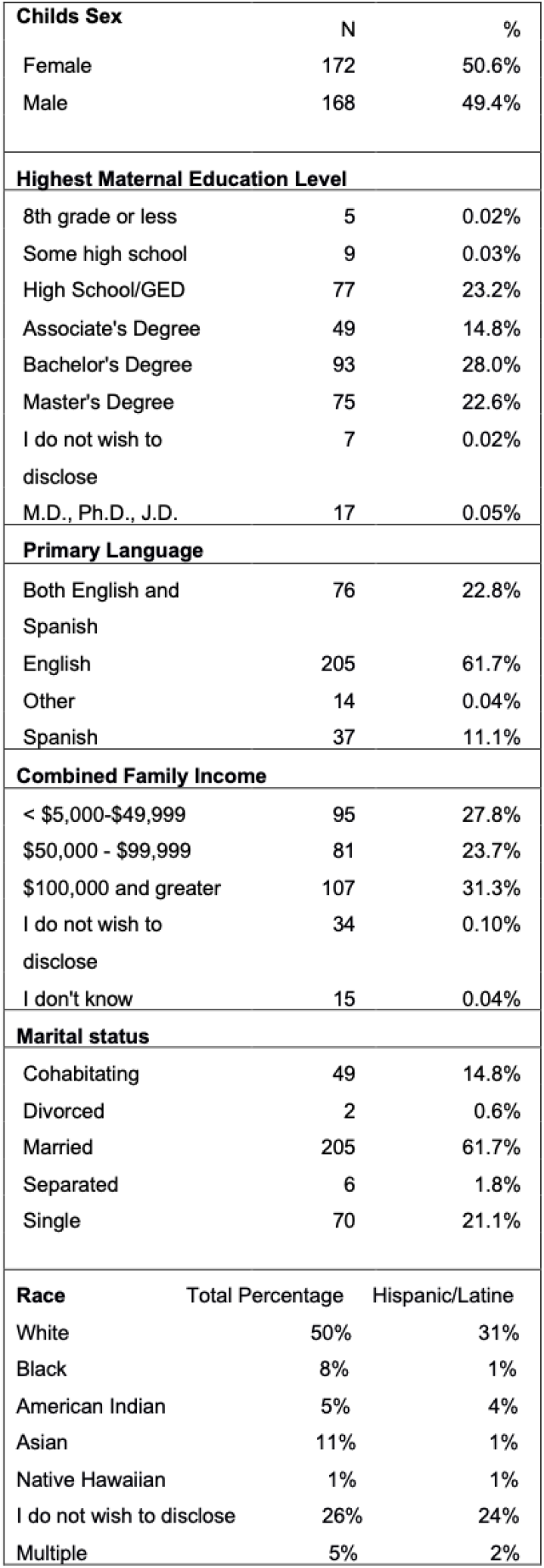
Distribution and means of sociodemographic characteristics of study participants.

### Adversity, Stress, Mental Health, and Development

Descriptive statistics of adversity and maternal emotional state are presented in Table 2. Maternal exposure to 4 or more ACES was reported by 28% of mothers (Table 2). Endorsements of at least 1 adverse experience in children (PEARLS Part 1) ranged between 36-39% of participants across the 3 study time points (Table 2). At 6 months, maternal PHQ-9 (M = 3.3, SD = 3.9) and PSS (M = 12.1, SD = 6.5) scores indicated generally low levels of depressive symptoms and perceived stress. These trends remained consistent at 12 and 24 months. QUIC 5 scores at 12 and 24 months were low on average (M = 0.4, SD = 0.7; M = 0.37, SD = 0.73), respectively (Table 2), indicating relatively stable home environments. Mean Bayley 4 cognitive, language, and motor standard scores are also presented in Table 2. Between 6-24 months Cognitive scores saw a general decrease, from 110 (95% CI 109,111) at 6 months, to 103 (95% CI 101,106), yet remained relatively stable at 24 months (Table 2). A similar trend was seen for language and motor scores between 6- and 24-months.

**Table 2.** Descriptive statistics of Adversity and mental health questionnaire’s IsoPs and child development scores. **Change from 6-24 months evaluated by linear mixed effects model (β= -0.4,p=<0 001) *Change from 6-24 months evaluated by linear mixed effects model (β= -0.2,p=003)

| Measure |  | 6 months (n=340) | 12 Months (n=292) | 24 Months (n=119) |
| --- | --- | --- | --- | --- |
| **Questionnaires** |  |  |  |  |
| Adut ACES |  |  |  |  |
|  | 0 | N=116 (34%) | N/A | N/A |
|  | 1-3 | N=139 (41%) | N/A | N/A |
|  | 4+ | N=85 (25%) | N/A | N/A |
| PEARLS Pt1 |  |  |  |  |
|  | 0 | N=220 (65%) | N=192 (66%) | N=78(66%) |
|  | 1+ | N=120 (35%) | N=100 (34%%) | N=41(34%) |
| **Mean (SD)** |  |  |  |  |
| QUIC 5 |  | N/A | 0.4 (0.7) | 0.37 (0.73) |
| PHQ9 |  | 3.3 (3.9) | 3.5 (4.3) | 3.4 (4.5) |
| PSS |  | 12.1 (6.5) | 12.5 (6.0) | 11.3 (6.3) |
| **F2-Isoprostaines (ng/mgCr)** |  |  |  |  |
| 15-F<sub>2t</sub>-IsoP |  | 2.4 (1.9) | 2.2 (2.2) | 1.4 (1.4)** |
| 5-F<sub>2t</sub>-IsoP |  | 4.6 (4.5) | 4.6 (5.0) | 3.5 (2.7)* |
| **Bayley-4** |  |  |  |  |
| **Mean (95% CI)** |  |  |  |  |
| Cognitive Standard Score |  | 110 (109,111) | 103 (102,105) | 104 (101,106) |
| Language Standard Score |  | 105 (105,106) | 99 (98,101) | 100 (96,104) |
| Motor Standard Score |  | 109 (108,110) | 104 (103,106) | 105 (102,107) |

### Biomarkers

F2-isoprostane levels exhibited a general decline over time. Mean 15-F_2t_-IsoP levels decreased significantly from 2.4 ng/mgCr at 6 months to 1.4 ng/mgCr at 24 months (β=-0.4,p=<0.001). Similar declines were noted for 15-F_2t_-IsoP from 4.6 to 3.5 ng/mgCr (β=-0.2,p=0.003) (Table 2).

### Correlations Among Adversity, Biomarkers, and Development

Time-lagged analyses indicated that higher maternal ACEs scores at 6 months were positively correlated with higher 5-F_2t_-IsoP (r=0.15, p=0.055) and 15-F_2t_-IsoP (r = 0.24, p = 0.02) (Figure 1A,B) at 12 months. Higher 12-month QUIC 5 score correlated with higher 24 month 5-F_2t_-IsoP (r=0.43, p=0.05)(Figure 1C).

**Fig 1.**
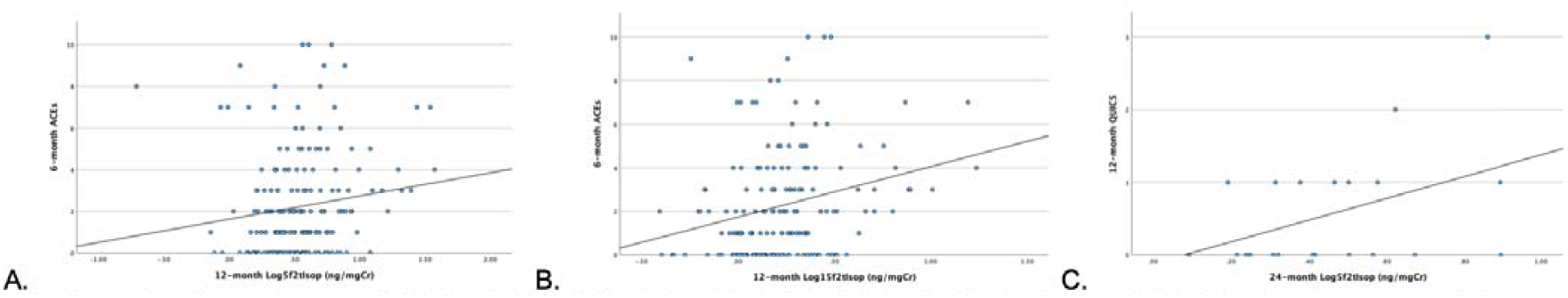
Scatter plot of 6-month maternal ACES and 5-F_2t_-lsoP (r=0.15,p=0.055) (A) and 15-F_2t_-lsoP (r = 0.24, p = 0.02)(B).QUIC-5 at 12 months correlation to 5-F_2t-_IsoP at 24-months (r=0.43, p=0.05) (C).

A PEARLS endorsement was not correlated significantly with F2_α_ -IsoPs. Cross-sectional analyses demonstrated significant correlations at 6 months between higher 15-F_2t_-IsoP levels and lower Bayley language (r = -0.15, p = 0.02) and motor (r = -0.16, p = 0.02) scores (Figure 2A & 2B), but not at later timepoints.

**Fig 2.**
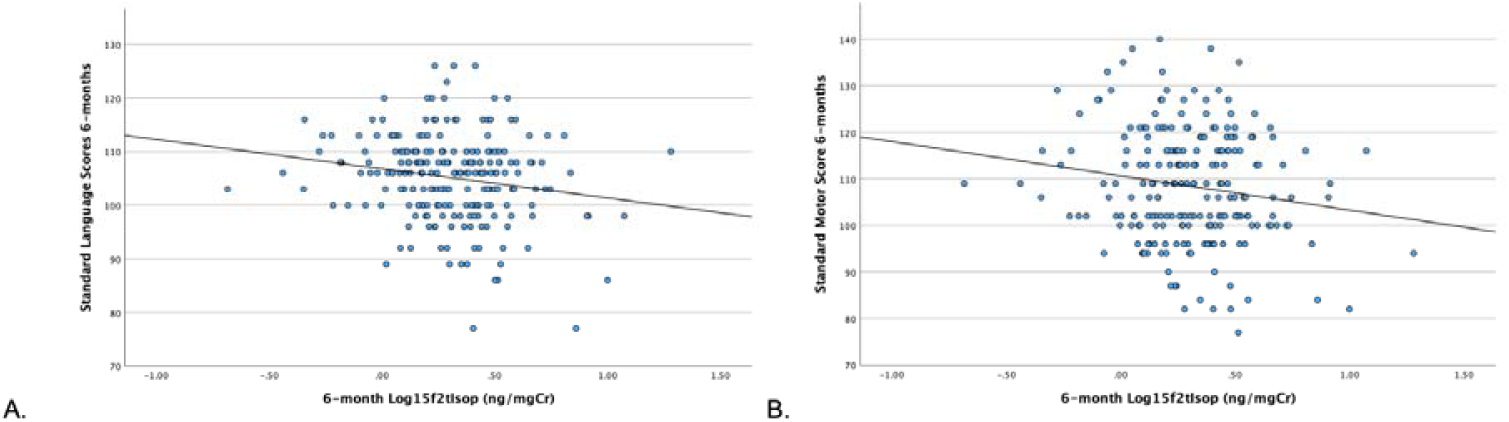
Scatter 6-month Bayley-4 language score (A) (r = -0.15. p = 0.02) and 6-month Bayley-4 motor score (B) (r = -0.16. p = 0.02) and 15-F_2t-_lsoP at 6 months.

### Modeling F_2α_-IsoPs Relation to Maternal Adversity

Spaghetti plots demonstrating the change in 15-F_2t_-IsoP and 5-F_2t_-IsoP from initial study timepoint at 6 months to 24 months are depicted in Figure 3.

**Fig 3.**
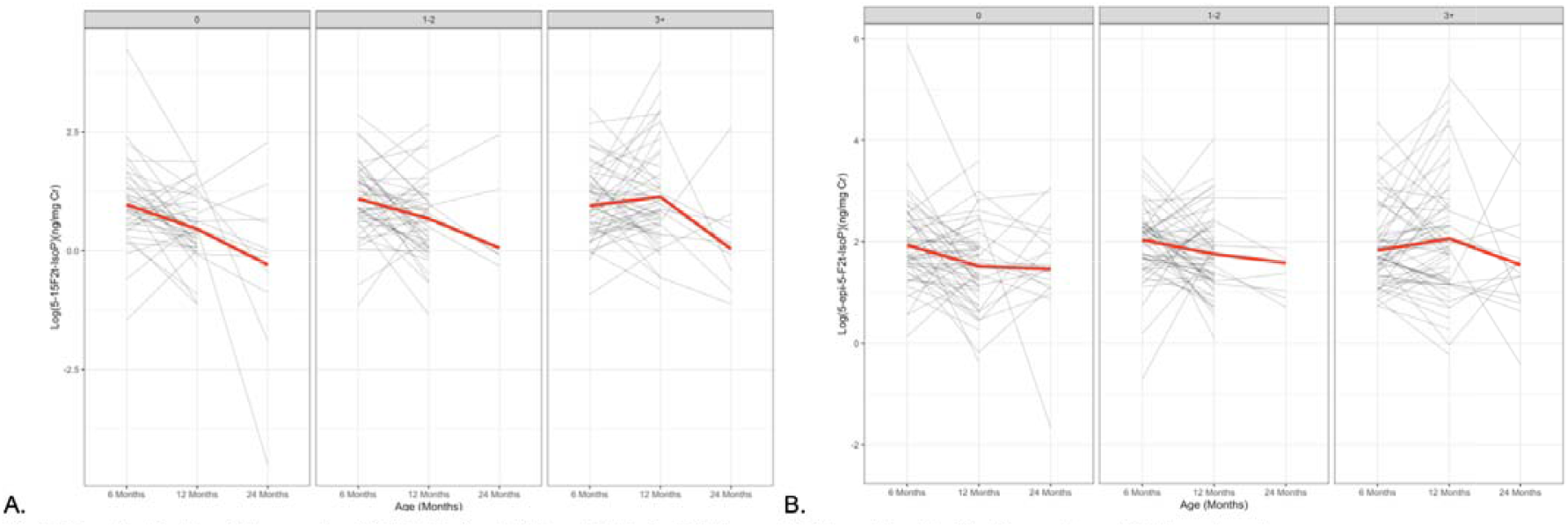
Spaghetti plots of change in child 15-F_2t_-lsoP (A) and 5-F_2t-_lsoP (B) over 6-24 months stratified by maternal ACEs categories.

To evaluate how the change in 15-F_2t_-IsoP and 5-F_2t_-IsoP over time varied by maternal ACES scores, a piecewise linear aggression was done (Table 3). There was a decline in 15-F_2t_-IsoP and 5-F_2t_-IsoP levels between 6-12 months in children whose mothers had an ACEs score of 0 (Table 3). In children whose mothers had ACES scores ranging from 1-2, the decrease in 15-F_2t_-IsoP and 5-F_2t_-IsoP between 6-12 months was less than in children whose mothers had 0 ACEs (Table 3). In contrast to the declines, when evaluating children whose mothers had ACEs scores 3 or greater, there was an increase in 15-F_2t_-IsoP levels between 6-12 months (Table 3). When comparing the change in F_2α_-IsoPs levels across the three different groups of maternal ACEs exposure, a history of maternal adversity impacted the change in 15-F_2t_-IsoP (p=0.003) and 5-F_2t_-IsoP(p=0.003) levels between 6-12 months (Table 3).

**Table 3.**
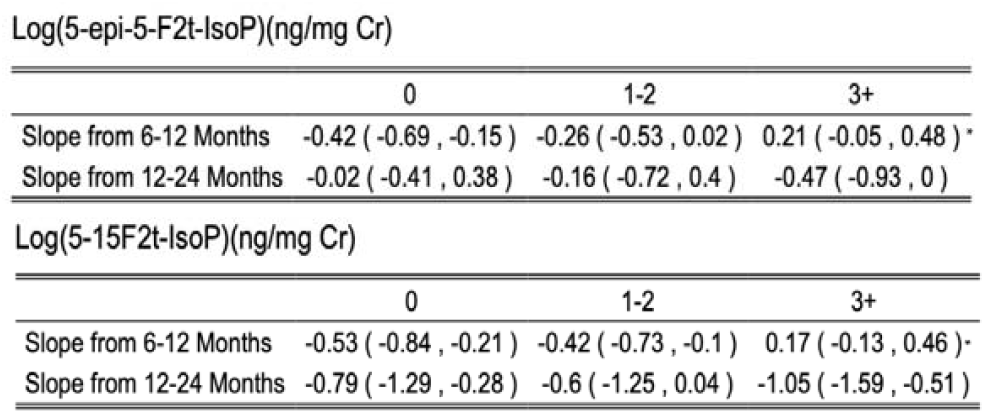
Piecewise linear analysis evaluating change In 15-F_2t-_lsoP and 5-F_2t-_lsoP between 6-12 months and 12 to 24 months by maternal ACES category. β values and 95% confidence intervals displayed. *p-value=0.003, p-values represent that the slope for each ACEs group are significantly different from one another.

Lastly, when comparing the mean difference in F_2t_-IsoP levels over 6-12 months amongst different maternal ACEs categories, infants with maternal ACEs score of 0 vs 3+ and 1-2 vs 3+ showed a significant mean difference for both 15-F_2t_-IsoP and 5-F_2t_-IsoP (Table 4).

**Table 4.**
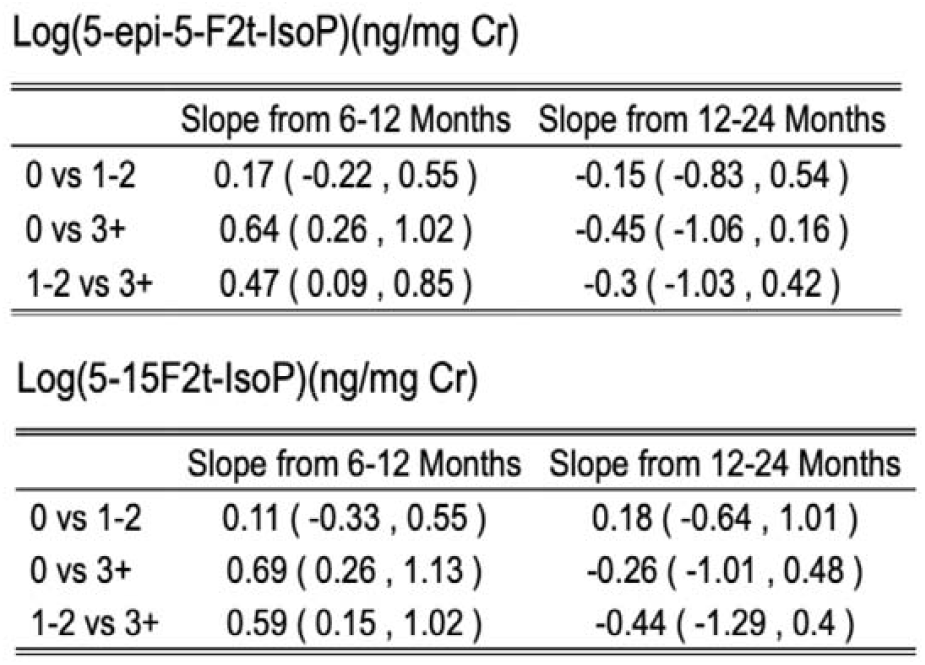
Mean difference in slopes in F_2t-_lsoP across maternal ACEs categories. Mean difference of β and 95% confidence intervals represented.

## Discussion

The present analysis of an ongoing longitudinal study of 340 maternal-child dyads, representing the demographics of the greater Los Angeles basin, evaluated the possible relations between maternal ELA and infant oxidative stress and neurodevelopmental outcomes over the first two years of childhood. We found that maternal adversity correlated with higher oxidative stress in infants at 12 months. We also found that higher infant oxidative stress correlated with lower neurodevelopmental scores at the 6-month timepoint. An additional novel finding is that the change in infant oxidative stress over the first year of life demonstrated that maternal ELA, captured by the ACEs questionnaire, was associated with F_2α_-IsoPs trajectory in a dose dependent manner. The findings suggest that an oxidative stress state in early development identifies a subset of infants impacted by environmental factors that may have physiologic and neurodevelopmental consequences. It is important to note that while maternal adversity correlated with 12-month F_2α_-IsoP, there was an earlier correlation between elevated infant F_2α_-IsoPs at 6 months with lower neurodevelopment at the same age. Though these relations help inform a physiologic pathway and window for understanding how oxidative stress impacts development, these findings represent a cross section in time. More informative relations come from understanding changes in oxidative stress over time as it relates to ELA and neurodevelopment.

Identifying biomarkers as targeted, personalized, physiologic measures of psychosocial experience as early as possible is the next frontier in ELA research. Given the dynamic nature of development and how critical periods inform long-term health outcomes, gaining new insight on the impact of oxidative stress on development over time is crucial for improving clinical translation of screening methods. Experts in the field of ELA have set biomarker identification as a critical need(18). In our previous exploratory study of 110 maternal infant dyads, we found that maternal perinatal cumulative risk (inclusive of socioeconomic disadvantage and poorer mental health) correlated with elevated maternal and infant 15-F_2t_-IsoP at 6 months(58). In the current longitudinal study with a new cohort of 340 maternal-child dyads, the analyses revealed that higher prenatal maternal ACEs is correlated with elevated infant 15-F_2t_-IsoP and 5-F_2t_-IsoP at the later time point of 12-months. We also found that higher household unpredictability was associated with elevated child 5-F_2t_-IsoP at 24 months. These findings build upon our previous study and suggest that there may be an impact of prenatal maternal adversity and subsequent postpartum home instability on infant oxidative stress status that extends into the second year postnatally. Additionally, household unpredictability has been linked to psychopathology and physical health problems in adolescents. The current study is the first to evaluate household unpredictability in relation to early childhood oxidative stress(4, 64).The association between maternal prenatal/perinatal ELA and maternal oxidative stress has been identified in previous studies(71-74). Here, we establish the relation between maternal adversity and toddler oxidative stress, which had not been fully determined previously. The importance of identifying risk factors that may inform later child outcomes presents a unique opportunity for maternal interventions prior to pregnancy and birth.

While prior studies have identified a relationship between prenatal/perinatal ELA and altered neurodevelopment in children, the biologic pathway has yet to be fully established(9, 75). Previously, we proposed that oxidative stress mediated the relationship between perinatal adversity and altered infant neurodevelopment(58). We reported that infant oxidative stress at 2 months predicted lower neurodevelopmental scores at 12 months(58). In the current study we found that 6-month-old infants with elevated 15-F_2t_-IsoP levels had lower language and motor scores. While previous studies have reported relations between maternal oxidative stress and infant development, our current and previous work demonstrating associations between child oxidative stress and subsequent neurodevelopment provides further evidence for the involvement of energetics pathways(76, 77).

Developmental changes over time are important in considering how early status relates to later outcomes. Thus, we characterized infant 15-F_2t_-IsoP and 5-F_2t_-IsoP levels for changes over time. The data revealed that on average, when including all infant participants, F_2α_-IsoPs levels decreased from 6-24 months. Research by Freil et al. found that at birth, infants have high oxidative stress as a result of transition from in utero hypoxia to relative hyperoxia and that that oxidative stress rapidly declines over the first 6 months, when F_2α_-IsoPs levels are similar to those in adults(78). When evaluating F_2α_-IsoPs trajectory by maternal perinatal adversity categories: 0,1-2, and 3 or more, the expected F_2α_-IsoPs decrease was evident in infants with no maternal adversity history. However, in infants whose mothers endorsed 1-2 ACEs, the F_2α_-IsoPs decline was less than what was seen in infants with no maternal ACEs history. A key finding of the present study is that infants whose mothers endorsed a history of 3 or more ACEs, there was an increase, rather than the expected decrease, in 15-F_2t_-IsoP and 5-F_2α_-IsoPs levels between 6-12 months. Regression analysis revealed statistically significant differences between the three different trajectories across ACEs categories. These findings have important implications, noting that averaging datasets may fail to reveal important subgroups that exhibit distinct oxidative stress responses to ELA and maternal care. The 12-24-month trajectories for all ACEs categories demonstrated a decline in levels with the three trajectories not being significantly different. This in part may be due to lower sample sizes at the later study timepoints that are currently on going.

There are several limitations to the current study. Given that this is a longitudinal study in which later study time points have yet to be completed, we interpret the results of those time points with an abundance of caution and recognize final relations may vary based on attrition and development. Additionally, it is unlikely that a single biomarker will be sufficient in identifying children with a toxic stress response. Thus, other measures of mitochondrial health will be necessary to fully elucidate the biological mechanisms underlying the toxic stress response. Those studies are underway with several other biomarkers of mitochondrial health to be described in subsequent publications. The questionnaires used to assess ACEs and maternal emotional state evaluate narrow dimensions of adversity. More robust and comprehensive measures assessing experiences of adversity including but not limited to racism/discrimination, socioeconomic disadvantage, impacts of displacement or immigration are needed to more wholistically quantify experiences of adversity. The F_2α_-IsoPs regiosomers evaluated in this study, while the most prevalent, are not inclusive of all regiosomers representative of oxidative stress. Additionally, recent studies have found that metabolites of the commonly studied regiosomers may add novel information regarding health outcomes(79, 80).

## Conclusions

With the above limitations noted, we conclude that from this high ACEs endorsing, modest sized cohort, important relations between prenatal maternal adversity, oxidative stress, and alterations in child neurodevelopment, as well as child oxidative stress status, will be important to consider in future studies of toxic stress response.

## Data Availability

The datasets used and/or analyzed during the current study are available from the corresponding author on reasonable request

## Declarations

### Ethics approval and consent to participate

This study was approved by the Children’s Hospital Los Angeles Institutional Review Board. Informed consent was obtained from all adults participating in the study and by parent or guardian for all minor participants. This study adhered to the Declaration of Helsinki. Clinical trial number: not applicable

### Consent for publication

Not applicable

### Competing Interests

The authors declare that they have no competing interests.

### Funding

This work is supported by The California Initiative to Advance Precision Medicine, The Treehouse Family Foundation, and TSRI Developmental Neuroscience and Neurogenetics Program.

### Authors contributions

KG made substantial contributions to the acquisition, analysis and interpretation of the data, drafted and revised manuscript draft. RDA made substantial contributions to the analysis and interpretation of the data and revisions of the manuscript. PL made substantial contributions to the conception and design of the study, interpretation of the data and substantial revisions of the manuscript.

## Abbreviations

ELA: Early life adversity
ACE: Adverse childhood experience
F_2α_-IsoPs: F_2α_-Isoprostanes
ROS: Reactive oxygen species
PEARLS: Pediatric ACEs and related life events screener

## Acknowledgements

We would like to acknowledge the research associates Dianna Guerrero Jimenez, Aime Ozuna, Nicole Fonacier, Evelyn Romo, and Jenny Legido for their dedication to excellence in data collection and biospecimen processing.

## Notes

### Competing Interest Statement

The authors have declared no competing interest.

### Author Declarations

This study was approved by the Childrens Hospital Los Angeles Institutional Review Board. Informed consent was obtained from all adults participating in the study and by parent or guardian for all minor participants. This study adhered to the Declaration of Helsinki.

